# Gait initiation impairment in patients with Parkinson’s disease and freezing of gait

**DOI:** 10.1101/2022.08.26.22279211

**Authors:** Chiara Palmisano, Laura Beccaria, Stefan Haufe, Jens Volkmann, Gianni Pezzoli, Ioannis U. Isaias

## Abstract

**Objective:** Freezing of gait (FOG) is a sudden episodic inability to produce effective stepping despite the intention to walk. It typically occurs during gait initiation or modulation and may lead to falls and loss of independence. We assessed gait initiation (GI) changes in parkinsonian patients suffering from FOG when off dopaminergic medications, removing correlations with anthropometric measurements, the base of support, and initial stance posture. We also addressed the temporal pattern of segmental body movements subserving gait initiation.

**Methods:** We studied 23 subjects with Parkinson’s disease (PD) and FOG (PDF), 20 patients with PD and no previous history of FOG (PDNF), and 20 healthy controls (HC). Kinematic and dynamic analysis of anticipatory postural adjustments (imbalance, unloading, stepping phase) was performed while patients were starting gait from upright standing after a self-selected period, to avoid any effect of cueing on gait initiation.

**Results:** The center of pressure (CoP) displacement and velocity during imbalance showed a significant impairment in both PDNF and PDF, more prominent in the latter patients. Several measurements were specifically impaired in PDF patients, especially the CoP displacement along the anteroposterior axis during unloading. The pattern of segmental center of mass (SCoM) movements during gait initiation did not show any differences between groups, but had high inter-subject variability. The standing postural profile preceding GI did not correlate with outcome measurements.

**Conclusions:** Our results showed impaired motor programming at gait initiation in parkinsonian patients. The more prominent deterioration of unloading in PDF patients might suggest impaired processing and integration of somatosensory information subserving gait initiation. The unaltered temporal movement sequencing of SCoM in both PDNF and PDF might indicate some compensatory cerebellar mechanisms triggering time-locked models of body mechanics in PD.

## 1 Introduction

Freezing of gait (FOG) is a dramatic phenomenon frequently affecting patients with Parkinson’s disease (PD) (Perez-Lloret et al., 2014), causing falls, mobility restrictions, and poor quality of life (Kerr et al., 2010; Okada 2011; Pelykh 2015). FOG is defined as a brief, episodic absence or marked reduction of forward progression of the feet despite the intention to walk (Nutt et al., 2011), which typically occurs when initiating or modulating gait (e.g., turning, obstacle crossing, etc.).

Gait initiation (GI) is a highly challenging task for the balance control system, and is of particular interest in the study of neural control of upright posture maintenance during whole-body movement (Yiou et al., 2017). Specifically, this task allows the precise assessment of anticipatory postural adjustments (APAs; i.e., muscular synergies that precede GI), aiming to destabilize the antigravitary postural set by shifting the center of pressure (CoP) to generate a gravitational moment favoring the center of mass (CoM) forward acceleration (Crenna and Frigo, 1991). The APAs are considered a motor program controlled by feedforward mechanisms regulated by the supraspinal locomotor network (Petersen et al., 2001; Hiraoka et al., 2006; Jacobs et al., 2009a; Palmisano et al., 2020a,b). The selection and scaling of appropriate APAs rely on the ability to use sensory information to determine the body positioning relative to the environment prior to step execution (Inglis et al., 1994; Mouchnino et al., 2015) and on the intended forthcoming movement (natural, slow, fast, obstacle, etc.) (Brenière et al., 1987; Crenna and Frigo, 1991; Lepers and Brenière, 1995; Caderby et al., 2014).

Striatal dopamine loss, a pathophysiological hallmark of PD, greatly impacts the production of APAs at GI and particularly the CoP displacement and velocity (Palmisano et al., 2020a). Only few studies have specifically investigated the GI task in parkinsonian patients with a history of FOG (PDF), non-implanted for deep brain stimulation (DBS) and after withdrawal of dopaminergic medication (meds-off state). The stimulation and medication condition should be carefully considered, as both DBS and dopaminergic drugs can variably influence posture and gait in PD (Burleigh-Jacobs et al., 1997; Frank et al., 2000; Crenna et al., 2006; Liu et al., 2006; Rocchi et al., 2006; Chastan et al., 2009; Pötter-Nerger and Volkmann, 2013; Mazzone et al., 2014; Curtze et al., 2015; Palmisano et al., 2020a). Overall, these studies showed conflicting results, with APAs being reported as normal (Nonnekes et al., 2014; de Souza Fortaleza et al., 2017; Schlenstedt et al., 2018) or multiple and hypometric (Jacobs et al., 2009b; Cohen et al., 2017). Several methodological discrepancies may account for such different findings, including a non-standardized meds-off state (Nonnekes et al., 2014), imposed (predefined) feet positioning (Schlenstedt et al., 2018), cueing (Jacobs et al., 2009b; Cohen et al., 2017; Schlenstedt et al., 2018), and specific instructions on the execution of the GI task (e.g., to start walking as quickly as possible (Cohen et al., 2017; Bayot et al., 2021) or while performing a cognitive task (de Souza Fortaleza et al., 2017). All of these factors can significantly impact and alter APAs expression at GI. Specifically, a cued start signal influences motor programming towards normalization, especially in PDF (Burleigh-Jacobs et al., 1997; Hiraoka et al., 2006), similar to the improvements seen with the administration of levodopa for self-generated step initiation (Burleigh-Jacobs et al., 1997). Also, the initial feet position (Rocchi et al., 2006; Dalton et al., 2011; Palmisano et al., 2020a) and posture (Leteneur et al., 2013; Fortin et al., 2015; Fawver et al., 2018) can significantly impact the biomechanical features of APAs at GI.

Postural changes in particular would have a detrimental impact on APAs production. An altered representation of the body position (egocentric representation) may determine a functional re-organization of the supplementary motor area (SMA)-proper hampering selection and re-scaling of APAs to adapt to the altered postural framework and bradykinetic stepping (Leteneur et al., 2013; Fortin et al., 2015; Yoshii et al., 2016; Delafontaine et al., 2017; Stansfield et al., 2018).

This study aims to describe GI alteration in patients with PD and FOG accounting for the influence of anthropometric measurements (AM) and the base of support (BoS) and investigating their relationship with the initial posture. We have also addressed the relative timing and movement sequence of each body segment subserving GI.

## 2 Methods

### 2.1 Subjects

We recruited 23 patients with idiopathic PD (according to the UK Brain Bank criteria) and an unambiguous, previous history of FOG (PDF). In addition, 20 patients with PD and no previous history of FOG (PDNF) and 20 HC were also included. HC and PDNF patients were chosen to match in terms of demographic and clinical data with the PDF group. Subjects with neurological diseases other than PD, including cognitive decline (i.e., Mini-Mental State Examination score <27), vestibular disorders, and orthopedic impairments that could interfere with gait were excluded. Disease severity was evaluated with the Unified Parkinson’s Disease Rating Scale motor part (UPDRS-III).

The study was approved by the local investigational review board and conformed to the declaration of Helsinki. All subjects gave their written informed consent prior to participation.

### 2.2 Experimental protocol

Patients were investigated in practical meds-off state, i.e., in the morning after overnight withdrawal (>12 h) of all dopaminergic drugs.

Kinematic data were recorded using an optoelectronic system with six cameras (sampling rate 60 Hz, SMART 1.10, BTS, Italy) and a set of 29 markers placed on anatomical landmarks (temples, acromions, lateral humeral condyles, ulnar styloids, anterior superior iliac spines [ASIS], middle thighs, lateral femoral condyles, fibula heads, tibial anterior side, lateral malleoli, Achilles tendon insertion, fifth metatarsal heads, halluxes, the seventh cervical vertebra [C7], point of maximum kyphosis, middle point between the posterior superior iliac spines [PSIS]) (Ferrari et al., 2008; Isaias et al., 2014). Eight additional technical markers were placed on the trochanters, the medial condyles, the medial malleoli and the first metatarsi for a short calibration trial which allowed the computation of the AM and BoS measurements (Palmisano et al., 2019, Palmisano et al., 2020a, Palmisano et al., 2020b). Markers traces were filtered with a 5th-order lowpass Butterworth filter (cut-off frequency: 10 Hz (Palmisano et al., 2019)). Dynamic measurements were recorded with a force plate working at a sampling rate of 960 Hz (KISTLER 9286A, Winterthur, Switzerland). The resulting signal was low-pass filtered (5th-order lowpass Butterworth filter) with a cut-off frequency of 30 Hz (Muniz et al., 2012; Palmisano et al., 2020b).

At the beginning of each trial, subjects stood upright on the force platform at a comfortable stance position for about 30 s. The initial stance position was not standardized, to prevent modification of the subject’s usual motor strategy to initiate gait (Palmisano et al., 2020a).

Participants were instructed to start walking after a self-selected period from a verbal signal, to avoid any effect of cueing on GI. The instruction given was: “Start walking at the moment of your choice”. Subjects were not instructed on the stepping leg to use, and they moved at their own pace until the end of the walkway. After a training session, at least three consecutive trials were recorded. The principal investigator supervised all participants during the experiment.

### 2.3 Biomechanical measurements

#### Anthropometric measurements (AM) and Base of Support (BoS)

For each subject, we measured the following AM (Table 1): body height, inter anterior-superior iliac spine distance, limb length, foot length, body mass, and body mass index. The AM were recorded over a period of 5 s of standing using eight additional markers, as described in Palmisano et al. (2020a). The AM were used for the estimation of the CoM of each body segment (SCoM), according to the anthropometric tables and regression equations proposed by Zatsiorsky (1998). For each trial, the base of support (BoS) area and BoS width were calculated. We also accounted for feet position asymmetry by measuring the foot alignment, the difference between feet extra-rotation angles, and the BoS opening angle (Palmisano et al., 2020a,b).

**Table 1.**
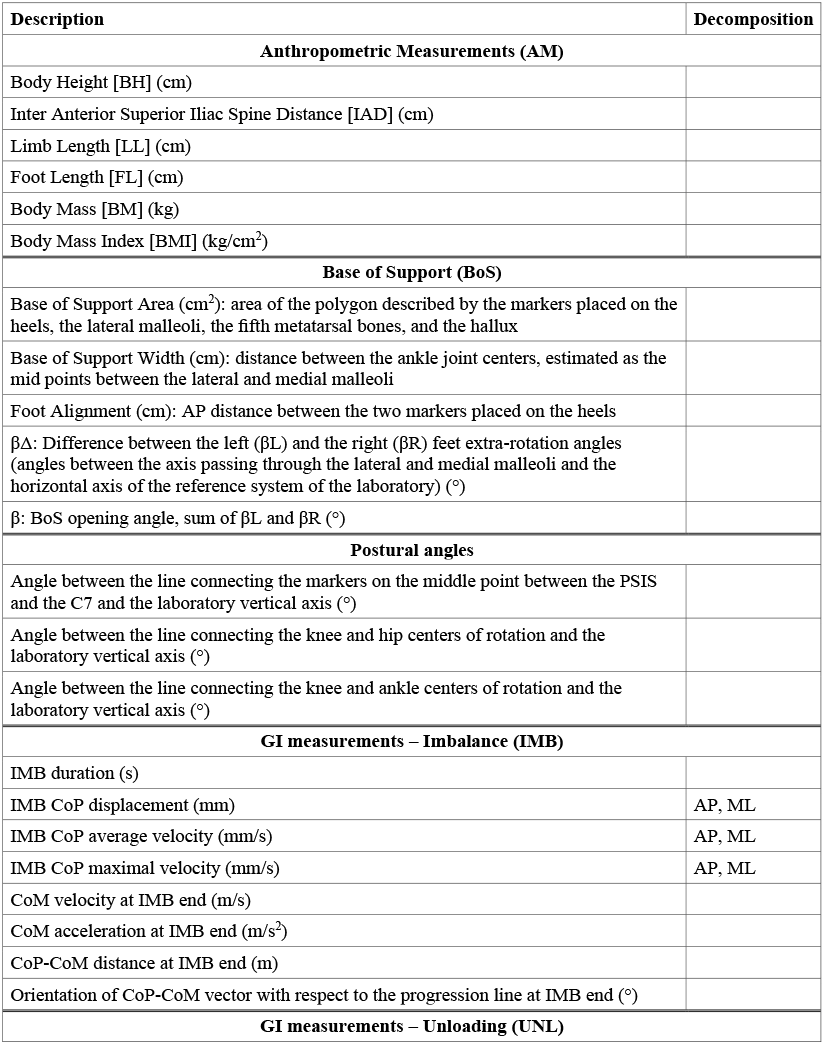
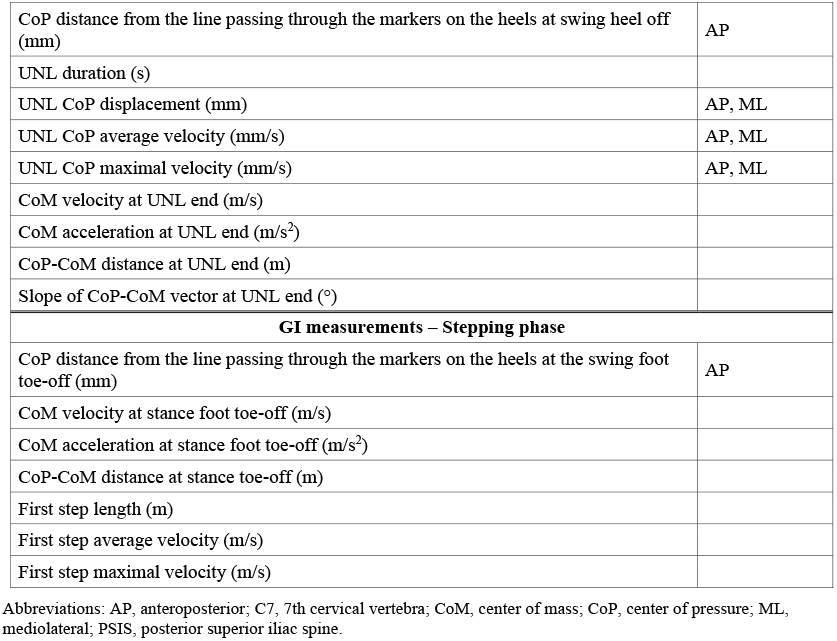
Biomechanical measurements.

#### Postural profile

The standing postural profile was characterized by means of trunk, thigh, and shank sagittal angles (Figure 1) (Crenna et al., 2006) computed shortly before the GI execution (during a 1-s window before the onset of the APAs). The trunk angle was defined as the inclination of the line passing through the markers placed on the middle point between the two posterior superior iliac spines and the 7th cervical vertebra with respect to the vertical axis of the laboratory. The thigh angle was calculated as the angle between the vector connecting the knee and hip center of rotation and the vertical axis of the laboratory. The shank angle was computed between the line connecting the joint centers of knee and ankle and the vertical axis of the laboratory.

**Figure 1.**
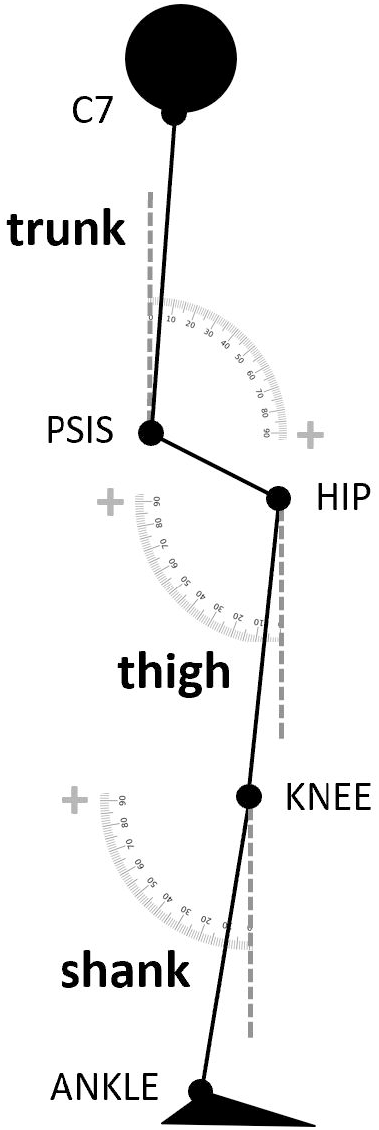
Postural angles. Scheme of the postural angles analysed in the study. The trunk angle was defined as the inclination of the line passing through the markers placed on the middle point between the two posterior superior iliac spines and the seventh cervical vertebra with respect to the vertical axis of the laboratory. The thigh angle was calculated as the angle between the vector connecting the knee and hip center of rotation and the vertical axis of the laboratory. The shank angle was computed between the line connecting the joint centers of knee and ankle and the vertical axis of the laboratory.

#### Anticipatory postural adjustments (APAs) and Gait initiation (GI)

GI variables were defined based on the displacement of the CoP, recorded by the force platform. The CoM was estimated as the weighted mean of the SCoM (Dipaola et al., 2016). GI variables were calculated by dedicated algorithms in Matlab ambient (Matlab® R2018b, The MathWorks Inc., Natick, MA, USA) (as in Palmisano et al., 2020a,b). All GI measurements computed in the study are listed and described in Table 1. Briefly, four reference instants were automatically identified on the CoP track and checked by visual inspection using an interactive software: the onset of the APAs, the heel-off of the swing foot (HO_SW_), the toe-off of the swing foot (TO_SW_) and the toe-off of the stance foot (TO_ST_). APAs onset (APA_ONSET_) was detected as the instant at which the CoP started moving consistently backward and toward the swing foot; HO_SW_ was defined as the time when CoP reached the most lateral position toward the swing foot; TO_SW_ was defined as the moment when the CoP shifted from lateral to anterior motion, and TO_ST_ as the last frame of the force platform signal (Figure 2). The APAs were divided into two periods: the imbalance phase (IMB), from APAs onset to HO_SW_, and the unloading phase (UNL), from HO_SW_ to TO_SW_ (Martin et al., 2002; Crenna et al., 2006; Isaias et al., 2014; Farinelli et al., 2020). The following measurements were calculated for both the IMB and UNL periods: duration and anteroposterior and mediolateral CoP displacement, average velocity and maximal velocity (Table 1). Of note, the mediolateral CoP displacement during the imbalance phase was considered positive when the shift of the CoP was towards the swing foot, while the mediolateral CoP displacement during the unloading phase was considered positive when the CoP was moving towards the stance foot. The IMB and UNL anteroposterior CoP displacement were both defined positive when the CoP movement was oriented backwards. We additionally defined the stepping phase, from HO_SW_ to the subsequent heel contact of the swing foot, by means of the markers placed on the feet. The first step was characterized in terms of step length and average and maximal velocity (Table 1). Velocity and acceleration of the CoM were defined at the end of the IMB and UNL phases and at the instant of TO_ST_. Additionally, the position of the CoM with respect to the CoP and the inclination of the vector connecting the two points in the transversal plane were computed at the end of IMB and UNL and at the TO_ST_ (Table 1) (Palmisano et al., 2020a).

**Figure 2.**
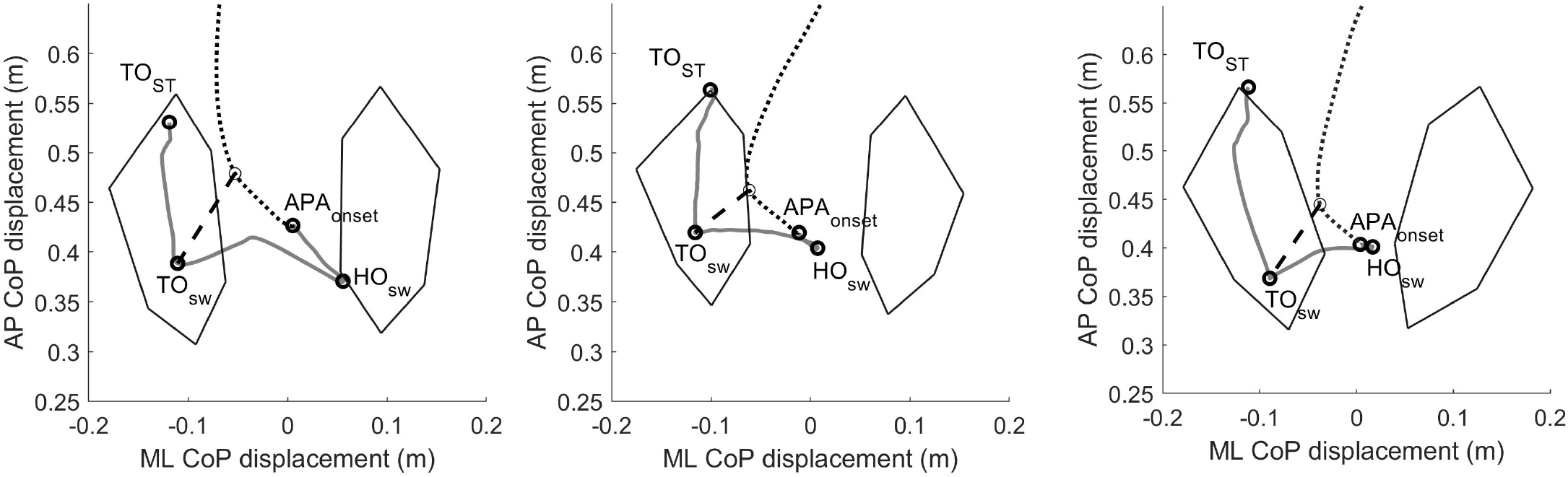
Two-dimensional center of pressure and center of mass trajectories during gait initiation. Example of the pathway of the center of pressure (CoP, grey solid line) and center of mass (CoM, black dotted line) during a gait initiation trial of one healthy subject (left panel), one parkinsonian patient without (PDNF, central panel) and one patient with (PDF, right panel) a positive history of freezing of gait. We defined the imbalance (IMB) phase as the interval between the onset of the APAs (APA_ONSET_) and the heel-off of the swing foot (HO_SW_), and the unloading phase (UNL) as the interval between the HO_SW_ and the toe-off of the swing foot (TO_SW_). The black dashed line represents the CoP-CoM vector at the end of the unloading (UNL) phase. With respect to healthy controls, the CoP displacement during the IMB phase was reduced for both PD and PDF patients. The CoP displacement during the UNL phase was in most of the cases backwards for the PDF patients only. Please see Table 3 for further details. Abbreviations: APAs, anticipatory postural adjustments; AP, anterior-posterior; CoP, center of pressure; HO, heel off; ML, mediolateral; TO, toe-off.

#### Segmental center of mass (SCoM)

To describe the temporal pattern of segmental movements during GI, we computed the latency of movement onset of the following 16 SCoM: head, chest, abdomen, pelvis, swing arm, stance arm, swing forearm, stance forearm, swing hand, stance hand, swing thigh, stance thigh, swing shank, stance shank, swing foot and stance foot (similarly to Rosin et al., 1997). For each trial, the movement onset latency of each SCoM was computed as the movement time from the onset of the APAs and normalized for the total GI time (from APA_ONSET_ to the toe-off of the swing foot). For each subject, we rank ordered the SCoM onset times and computed for each group: (i) the movement time from APA_ONSET_ normalized for the total GI time, and (ii) the relative frequency of each SCoM onset time to appear as events 1-16th of GI. To improve the readability of the data, we repeated the analysis after combining the SCoM into six groups (upper trunk: head and chest; lower trunk: abdomen and pelvis; swing arm: swing arm, forearm, and hand; stance arm: stance arm, forearm, and hand; swing leg: swing thigh, shank, and foot; stance leg: stance thigh, shank, and foot).

### 2.4 Statistical analysis

For each subject, all measurements were averaged over GI trials executed with the same swing foot. Each participant performed at least three GI trials with the same swing foot. Single trials and average values were inspected, and outliers removed from further analyses based on the Mahalanobis distance.

First, we verified the matching between groups for demographic, clinic, BoS and AM features with a Wilcoxon test (p-value set at 0.05). Before comparing the GI measurements across groups, we investigated their relationship with the BoS and AM with two partial correlation analyses (Palmisano et al., 2020a). For each group, we correlated the GI measurements first with the BoS measurements controlling for the AM, and then with the AM controlling for the BoS. In agreement with Palmisano et al. (2020a), GI variables that significantly correlated (Spearman’s ρ >0.5 and p-value <0.01) with the BoS in at least one group were excluded from further analyses. We opted for this conservative approach because the BoS was freely chosen by the subjects and may have been influenced by both the disease and compensatory mechanisms. The GI variables that correlated (Spearman’s ρ >0.5 and p-value <0.01) with the AM were instead corrected by means of the decorrelation normalization technique, as described in O’Malley (1996). This correction was applicable as AM were not influenced by the disease (no patient had camptocormia, skeletal deformities, etc.).

GI variables not dependent from the BoS and decorrelated from the influence of the AM were then compared between groups using a Dunn’s test (p-value set at 0.05, adjusted with Bonferroni correction for multiple comparisons).

We then investigated alterations of the initial postural condition. As for the GI measurements, we assessed the correlation of the AM and the BoS with the postural angles with partial correlation analyses (Spearman’s ρ >0.5 and p-value <0.01), before comparing the postural angles across groups (Dunn’s test, p-value set at 0.05, adjusted with Bonferroni correction for multiple comparisons).

As we found differences in the postural profiles across groups, we investigated if altered GI measurements in the PD groups were related to postural changes rather than to impaired motor programming. We performed a partial correlation analysis between the GI outcome measurements and the postural angles correcting for the group variable. We considered a correlation significant when Spearman’s ρ >0.5 and p-value <0.01.

Differences across groups in the SCoM movement onset were analyzed with a Dunn’s test (p-value <0.05, adjusted with Bonferroni correction for multiple comparisons).

All statistical analyses, except partial correlation analyses performed in Matlab, were performed with the JMP package (JMP® Pro 14.0.0, SAS Institute Inc., Cary, NC, USA).

## 3 Results

Demographic features, AM measurements, and BoS measurements did not significantly differ between groups (Table 2). Clinical data were similar between PDNF and PDF patients (Table 2).

**Table 2.**
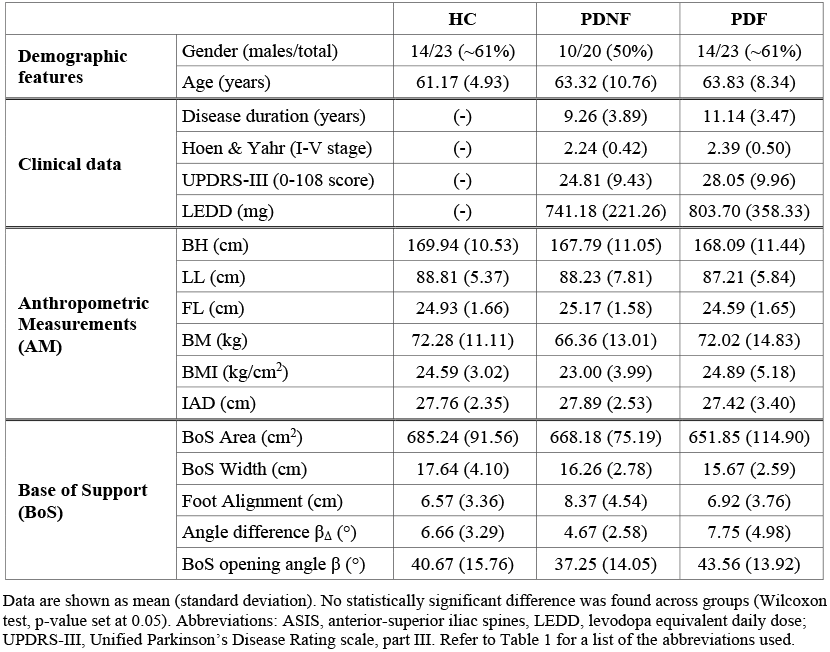
Demographic, clinical, anthropometric, and base of support features.

|  |  | **HC** | **PDNF** | **PDF** |
| --- | --- | --- | --- | --- |
| **Demographic features** | Gender (males/total) | 14/23 (~61%) | 10/20 (50%) | 14/23 (~61%) |
|  | Age (years) | 61.17 (4.93) | 63.32 (10.76) | 63.83 (8.34) |
| **Clinical data** | Disease duration (years) | (-) | 9.26 (3.89) | 11.14 (3.47) |
|  | Hoehn & Yahr (I-V stage) | (-) | 2.24 (0.42) | 2.39 (0.50) |
|  | UPDRS-III (0-108 score) | (-) | 24.81 (9.43) | 28.05 (9.96) |
|  | LEDD (mg) | (-) | 741.18 (221.26) | 803.70 (358.33) |
| **Anthropometric Measurements (AM)** | BH (cm) | 169.94 (10.53) | 167.79 (11.05) | 168.09 (11.44) |
|  | LL (cm) | 88.81 (5.37) | 88.23 (7.81) | 87.21 (5.84) |
|  | FL (cm) | 24.93 (1.66) | 25.17 (1.58) | 24.59 (1.65) |
|  | BM (kg) | 72.28 (11.11) | 66.36 (13.01) | 72.02 (14.83) |
|  | BMI (kg/cm<sup>2</sup>) | 24.59 (3.02) | 23.00 (3.99) | 24.89 (5.18) |
|  | IAD (cm) | 27.76 (2.35) | 27.89 (2.53) | 27.42 (3.40) |
| **Base of Support (BoS)** | BoS Area (cm<sup>2</sup>) | 685.24 (91.56) | 668.18 (75.19) | 651.85 (114.90) |
|  | BoS Width (cm) | 17.64 (4.10) | 16.26 (2.78) | 15.67 (2.59) |
|  | Foot Alignment (cm) | 6.57 (3.36) | 8.37 (4.54) | 6.92 (3.76) |
| | Angle difference $\beta_{\Delta}$ (°) | 6.66 (3.29) | 4.67 (2.58) | 7.75 (4.98) |
| | BoS opening angle $\beta$ (°) | 40.67 (15.76) | 37.25 (14.05) | 43.56 (13.92) |

Data are shown as mean (standard deviation). No statistically significant difference was found across groups (Wilcoxon test, p-value set at 0.05). Abbreviations: ASIS, anterior-superior iliac spines, LEDD, levodopa equivalent daily dose; UPDRS-III, Unified Parkinson's Disease Rating scale, part III. Refer to Table 1 for a list of the abbreviations used.

**Table 3.** Gait initiation measurements: comparison between groups.

|  | HC | PDNF | PDF |
| --- | --- | --- | --- |
| IMB duration (s) | 0.39 (0.08) | 0.38 (0.08) | 0.33 (0.09) |
| IMB displacement (mm) | 61.23<sup>+</sup> (20.32) | 35.54 (19.71) | 23.67<sup>+</sup> (9.94) |
| IMB displacement ML (mm) | 42.07<sup>#+</sup> (13.04) | 24.04<sup>#</sup> (13.61) | 17.76<sup>+</sup> (8.59) |
| IMB displacement AP (mm) | 36.53<sup>+</sup> (16.17) | 18.79 (14.75) | 9.16<sup>+</sup> (5.93) |
| IMB average velocity (mm/s) | 163.40<sup>#+</sup> (62.29) | 90.79<sup>#</sup> (47.29) | 84.03<sup>+</sup> (46.81) |
| IMB average velocity ML (mm/s) | 110.94<sup>#</sup> (34.90) | 62.14<sup>#</sup> (34.80) | 67.53 (41.47) |
| IMB average velocity AP (mm/s) | 103.36<sup>#+</sup> (53.78) | 47.74<sup>#</sup> (34.39) | 40.19<sup>+</sup> (30.45) |
| IMB maximal velocity (mm/s) | 344.22<sup>#+</sup> (149.41) | 189.88<sup>#</sup> (113.54) | 150.41<sup>+</sup> (64.26) |
| IMB maximal velocity ML (mm/s) | 238.29<sup>#+</sup> (77.82) | 137.25<sup>#</sup> (72.76) | 124.97<sup>+</sup> (56.96) |
| IMB maximal velocity AP (mm/s) | 225.81<sup>+</sup> (110.67) | 124.78 (65.52) | 101.41<sup>+</sup> (55.74) |
| IMB end CoM velocity (m/s) | 0.09<sup>+</sup> (0.03) | 0.06 (0.03) | 0.04<sup>+</sup> (0.02) |
| IMB end CoP-CoM distance (m) | 0.07<sup>+</sup> (0.02) | 0.04 (0.02) | 0.03<sup>+</sup> (0.01) |
| UNL duration (s) | 0.36 (0.08) | 0.40 (0.08) | 0.45 (0.19) |
| UNL displacement AP (mm) | -9.67<sup>+</sup> (15.30) | -6.25<sup>*</sup> (18.22) | 14.69<sup>+</sup> * (14.70) |
| UNL average velocity (mm/s) | 465.61 (162.21) | 323.94 (131.02) | 320.34 (150.20) |
| UNL average velocity ML (mm/s) | 422.79 (148.96) | 289.26 (121.24) | 290.67 (140.81) |
| UNL average velocity AP (mm/s) | 53.11 (20.97) | 37.29 (17.16) | 46.04 (34.21) |
| UNL maximal velocity AP (mm/s) | 344.48 (154.35) | 388.76 (169.88) | 359.19 (178.76) |
| UNL end CoM velocity (m/s) | 0.21<sup>+</sup> (0.06) | 0.16 (0.07) | 0.11<sup>+</sup> (0.04) |
| UNL end CoM acceleration (m/s<sup>2</sup>) | 1.29 (0.33) | 1.08 (0.41) | 1.12 (0.25) |
| UNL end CoP-CoM distance (m) | 0.08 (0.03) | 0.07 (0.03) | 0.07 (0.02) |
| ST toe-off CoM velocity (m/s) | 0.86<sup>#+</sup> (0.13) | 0.63<sup>#</sup> (0.24) | 0.53<sup>+</sup> (0.18) |
| ST toe-off CoM acceleration (m/s<sup>2</sup>) | 1.73<sup>+</sup> (0.38) | 1.28 (0.42) | 1.08<sup>+</sup> (0.33) |
| ST toe CoP-CoM distance (m) | 0.48 (0.32) | 0.51 (0.29) | 0.34 (0.28) |
| First step length (m) | 0.56<sup>+</sup> (0.07) | 0.43 (0.14) | 0.33<sup>+</sup> (0.13) |
| First step average velocity (m/s) | 0.98<sup>#+</sup> (0.14) | 0.68<sup>#</sup> (0.27) | 0.57<sup>+</sup> (0.27) |
| First step maximal velocity (m/s) | 3.21 (0.52) | 2.81 (0.72) | 2.64 (0.80) |

Of note, none of the patients showed freezing episodes during GI recordings. Therefore, our results define primarily the impact of APAs alterations and postural features in favoring FOG in Parkinson’s disease and not a causal correlation with the occurrence of gait freezing episodes at GI.

### 3.1 Selection of GI variables

The BoS did not correlate with most of the biomechanical measures of the IMB and stepping phases, but did correlate with the UNL. Results are consistent with our previous findings (Palmisano et al., 2020a). The GI variables that were independent from the BoS are listed in Table 3. The BoS and the AM showed no correlations with the trunk, thigh, and shank angles.

### 3.2 Postural features

The trunk and thigh angles, but not the shank angle, were significantly altered in both PDNF and PDF patients compared to HC (Table 4). Parkinsonian patients showed increased forward trunk bending associated with reduced thigh angle. The trunk was more flexed in PDF than PDNF patients, although this difference did not reach statistical significance. The thigh angle showed a negative average value only in the PDF group.

**Table 4.** Postural angles.

|  | **HC** | **PDNF** | **PDF** |
| --- | --- | --- | --- |
| Trunk (°) | 4.08<sup>#+</sup> (2.43) | 9.06<sup>#</sup> (4.37) | 12.58<sup>+</sup> (5.65) |
| Thigh (°) | 6.31<sup>#+</sup> (2.69) | 0.54<sup>#</sup> (3.70) | -0.48<sup>+</sup> (4.00) |
| Shank (°) | 9.22 (2.93) | 10.67 (2.77) | 10.95 (2.47) |

Postural angles were computed shortly before the GI execution (during a 1-s window before the onset of the APAs). Data are shown as mean (standard deviation). Dunn's test, significant p-value after Bonferroni correction: # HC vs. PDNF, + HC vs. PDF, \* PDNF vs. PDF.

### 3.3 Effect of PD and history of FOG on GI

We observed significant alterations in the GI execution in both PDNF and PDF patients, with the latter group showing overall more severely altered APAs measurements (Table 3, Figure 2).

The CoP displacement and velocity during IMB showed a progressive and significant reduction from HC to PDNF to PDF groups along both the mediolateral and anteroposterior axes.

The UNL and stepping phases were also altered in PDNF and PDF patients (Table 3, Figure 2). Of most relevance, in PDF the anteroposterior displacement of the CoP during UNL was backwards in most of the trials.

PDF patients showed significantly reduced first step length and both PD groups a lower first step average velocity compared to HC.

The CoM forward propulsion (velocity and acceleration) progressively decreased from HC to PDNF and to PDF.

### 3.4 Relationship between the standing postural profile and the GI

We did not find any significant correlation between the postural angles and the GI measurements. However, when not correcting for multiple comparisons, the shank angle was predictive for velocity variables of the IMB phase. Results are shown in Table 5.

**Table 5.** Correlation between the shank angle and gait initiation measurements.

|  |  | **Spearman's $\rho$** | **p-value** |
| --- | --- | --- | --- |
| Shank (°) | IMB average velocity (mm/s) | 0.32 | 0.014 |
|  | IMB average velocity AP (mm/s) | 0.31 | 0.016 |
|  | IMB maximal velocity AP (mm/s) | 0.38 | 0.003 |

Only significant partial correlations between postural angles and GI measurements corrected for the influence of the group variable are shown (Spearman's $\rho$ , p-value < 0.05). No correlation was significant after Bonferroni correction for multiple comparisons.

### 3.5 Pattern of movements during GI

The overall pattern of segmental movements during GI did not show clear differences between groups (Table 6 and 7). However, PDF showed lower times of movement onset for almost all ranked segments (Table 7), possibly suggesting tight inter-segmental coupling (Crenna et al., 2007). All groups started preferably with the swing or stance arm (Figure 3) especially the swing hand for HC and PDNF and the stance hand for PDF (Figure 4). The abdomen was often the last body segment moved by HC and PDNF, but not by PDF (Figure 4). Of note, we observed a remarkable inter-subject variability of SCoM onset times, especially for PD, as shown by the high value of the standard deviation (Table 6) and the large dispersion of the temporal order of SCoM movement onsets (Figure 3 and Figure 4), which probably prevented us to capture statistically significant differences.

**Table 6.** Onset of segmental movements at gait initiation.

| **Segmental center of mass (SCoM)** | **Movement onset** |  |  |
| --- | --- | --- | --- |
|  | **HC** | **PDNF** | **PDF** |
| Pelvis (%) | 60.62 (9.27) | 62.47 (13.74) | 62.66 (21.69) |
| Thigh ST (%) | 65.76 (10.75) | 71.72 (20.13) | 63.39 (19.97) |
| Shank ST (%) | 71.17 (14.36) | 68.57 (19.98) | 65.73 (21.95) |
| Foot ST (%) | 68.37 (10.46) | 77.24 (22.53) | 65.02 (22.87) |
| Thigh SW (%) | 62.67 (9.23) | 64.75 (15.47) | 55.85 (18.80) |
| Shank SW (%) | 74.21 (13.85) | 75.35 (24.26) | 67.26 (19.36) |
| Foot SW (%) | 69.15 (15.41) | 64.63 (16.55) | 59.20 (19.86) |
| Chest (%) | 73.48 (14.29) | 74.47 (26.12) | 69.22 (19.85) |
| Abdomen (%) | 79.50 (13.87) | 81.57 (25.30) | 57.88 (20.58) |
| Arm ST (%) | 55.49 (18.27) | 65.80 (26.63) | 54.17 (19.98) |
| Arm SW (%) | 62.71 (9.34) | 65.08 (13.49) | 56.57 (17.72) |
| Forearm ST (%) | 38.98 (12.65) | 53.73 (24.00) | 39.40 (12.69) |
| Forearm SW (%) | 51.34 (11.80) | 62.57 (13.59) | 47.26 (13.34) |
| Hand ST (%) | 44.37 (19.82) | 49.64 (21.32) | 41.99 (30.95) |
| Hand SW (%) | 34.36 (13.06) | 46.93 (19.96) | 47.71 (16.69) |
| Head (%) | 54.33 (11.81) | 58.15 (14.06) | 60.06 (18.41) |

Data are shown as mean (standard deviation). Time of movement onset of each segmental center of mass (SCoM) was expressed as the percentage with respect to total gait initiation duration (i.e., from the onset of the APAs to the heel contact of the swing foot) and compared across groups (Dunn's test, no difference was significant after Bonferroni correction for multiple comparisons).

**Table 7.**
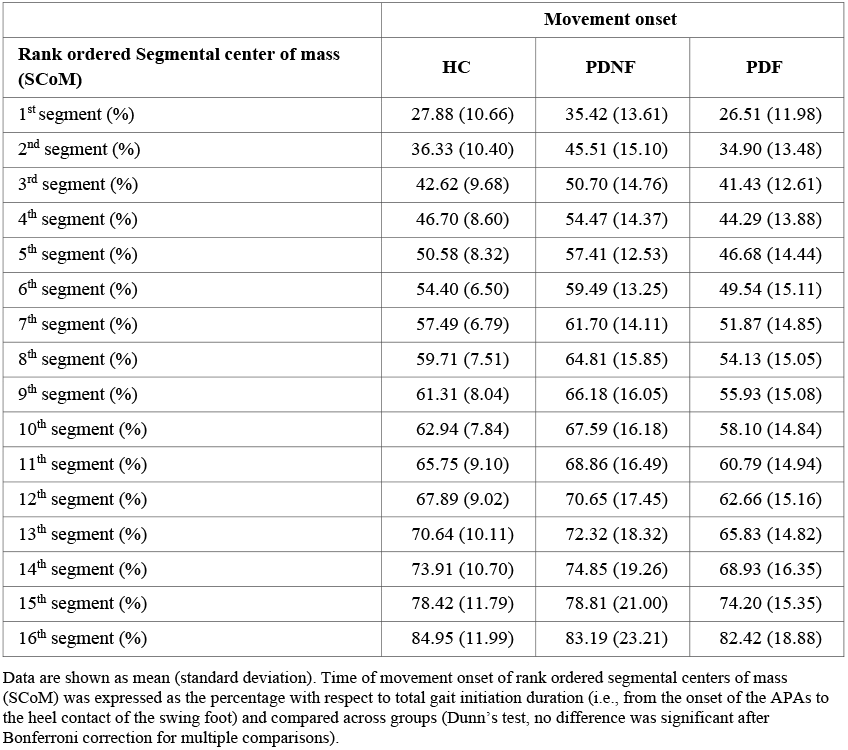
Onset of rank ordered segmental movements at gait initiation.

**Figure 3.**
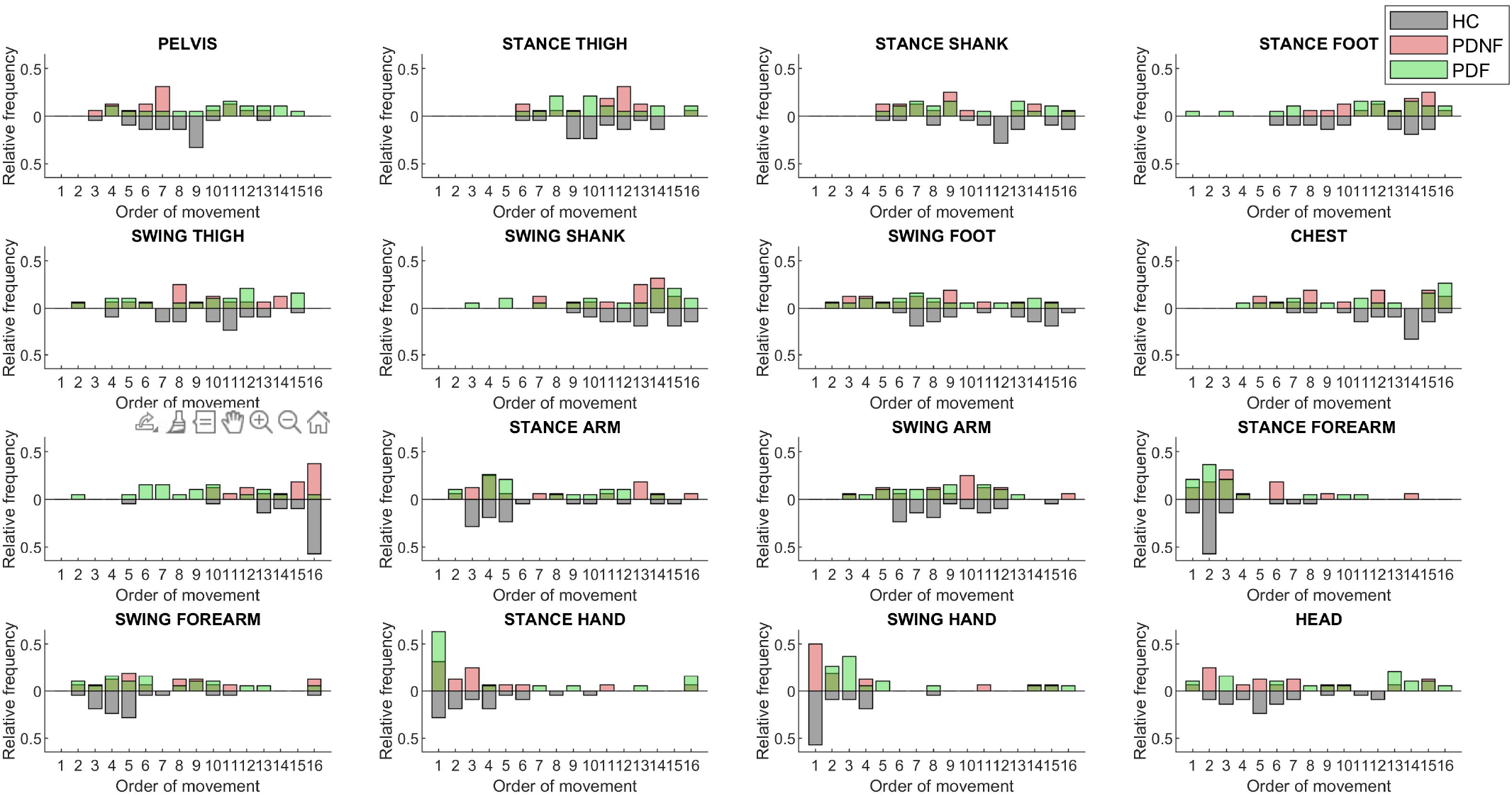
Rank order of Segmental CoM onset times. Temporal order of SCoM submovements within gait initiation for each group (grey: HC; red: PD; green: PDF). The bars represent the relative frequency with which each SCoM moves as 1-16^th^ segment along GI execution. Abbreviations: SCoM: segmental center of mass.

**Figure 4.**
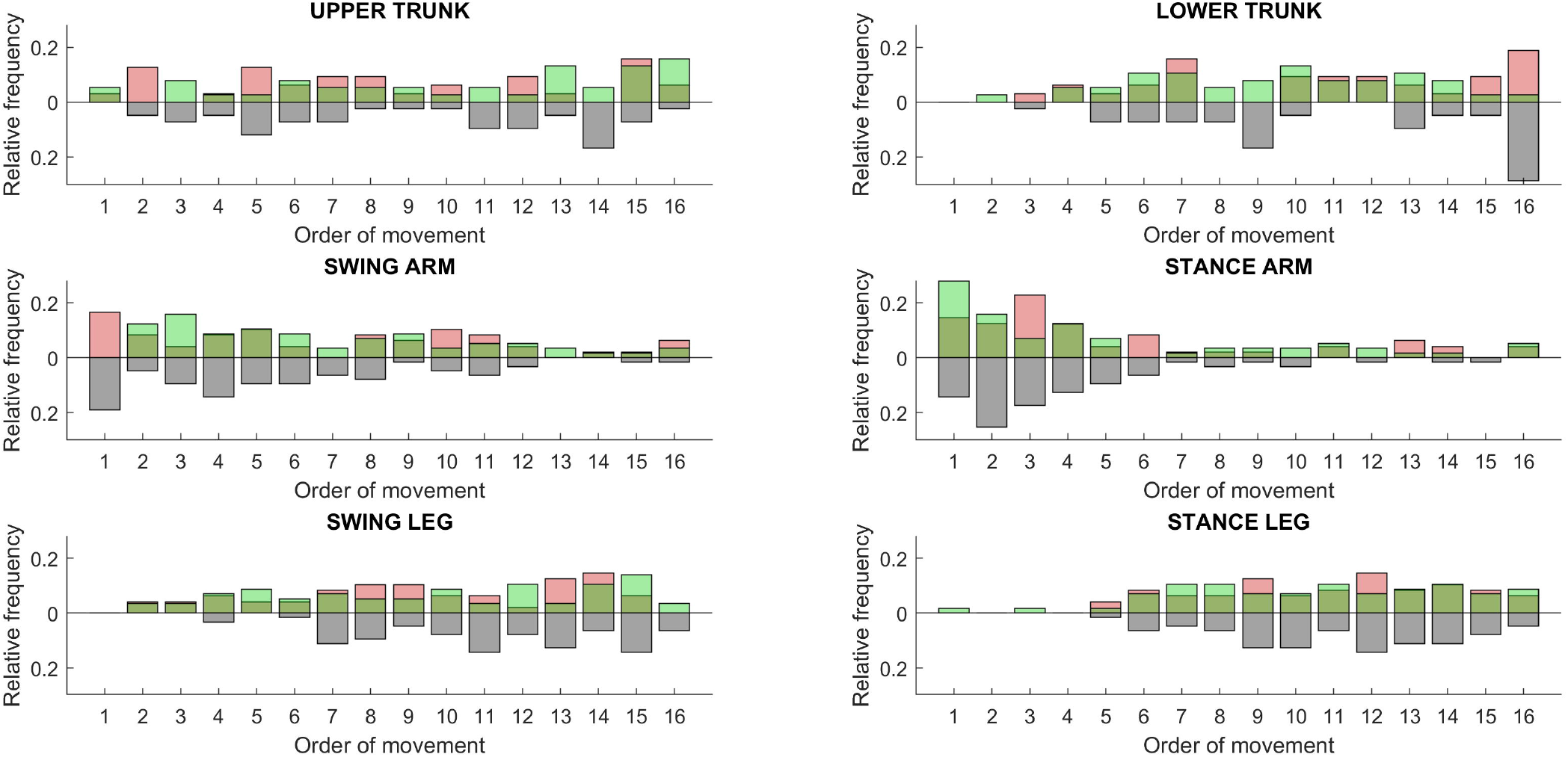
Rank order of onset times of groups of Segmental CoM. The rank ordered SCoM were classified into six groups (upper trunk: head and chest; lower trunk: abdomen and pelvis; swing arm: swing arm, forearm, and hand; stance arm: stance arm, forearm, and hand; swing leg: swing thigh, shank, and foot; stance leg: stance thigh, shank, and foot) and their temporal order of movement computed for each cohort (grey: HC; red: PD; green: PDF). The bars represent the relative frequency with which each SCoM moves as 1-16^th^ segment along GI execution. Abbreviations: SCoM: segmental center of mass.

## 4 Discussion

This study aimed to evaluate the specific biomechanical alterations of APAs at GI in PD patients with a positive history of FOG, accounting for known confounders such as medication condition, anthropometric measurements, base of support, and initial stance posture. Our findings are in line with previous studies in PD that showed an impairment in APAs production at GI (Halliday et al., 1998; Crenna et al., 2006). However, a direct comparison with earlier works is limited because as we aimed to minimize possible bias from cueing or imposed postural constraints that are known to affect the execution of the GI task (Burleigh-Jacobs et al., 1997; Dibble et al., 2004; Hiraoka et al., 2006; Rocchi et al., 2006; Plate et al., 2016; Schlenstedt et al., 2017; Palmisano et al., 2020a,b).

We have now shown that there is a profound alteration of APAs execution in PDF patients, which cannot be attributed to specific demographic or clinical features (such as disease severity and duration, medication dose and efficacy) as the patient groups were matched for all of these features (Heilbronn et al., 2019).

The IMB phase of APAs execution was significantly altered in all PD patients, particularly in PDF (Table 3). Increasing evidence suggests that this GI phase is governed by centrally-mediated feedforward signals and involves the cortico-basal ganglia loop, with the SMA-proper and the striatum chiefly contributing to the execution of these pre-programmed movements (Lee et al., 1999; Petersen et al., 2001; Hiraoka et al., 2006; Jacobs et al., 2009a; Bolzoni et al., 2015; Varghese et al., 2016; Richard et al., 2017; Yiou et al., 2017; Palmisano et al., 2020a,b). In PD, we have previously shown a detrimental effect of striatal dopamine loss in the IMB execution at GI (Palmisano et al., 2020a). Recent studies in parkinsonian patients suggested that striatal dopamine may in part enable normal movement by encoding sensitivity to the energy cost of a movement (Morris et al., 2006; Mazzoni et al., 2007; Schulz et al., 2007; Gepshtein et al., 2014). Therefore, from a perspective of motor planning, especially of patterned and consolidated motor actions such as APAs, a reduced tonic dopaminergic activity could reframe the coding of the expected energetic costs and impair motor control (Gepshtein et al., 2014).

In our study, we also showed a prominent alteration of the AP displacement of the CoP during the UNL phase in PDF patients. We interpret this result as a possible alteration, mainly of PDF patients, in the processing and integration of somatosensory information prior to stepping (Ruget et al., 2008; Mouchnino et al., 2014; You et al., 2017; Lhomond et al., 2019). A chief contribution to integrate proprioceptive and voluntary components for a proper weight transfer during GI can be expected from the premotor-parietal-cerebellar loop (Hanakawa et al., 1999; Picard and Strick, 2001; Voss et al., 2006; Wolbers et al., 2008; Bolzoni et al., 2015; Mouchnino et al., 2015; Tard et al., 2015). An impaired ability to inhibit stance postural control and initiate stepping and poor set-shifting are also included in pathophysiological hypotheses of FOG in PD (Jacobs and Horak, 2007; Amboni et al., 2008; Jacobs et al., 2009a,b; Naismith et al., 2010; Nutt et al., 2011; Heremans et al., 2013; Cohen et al., 2014; Lira et al., 2020).

Despite impaired APAs execution, the sequencing of the movement did not show major alterations in the PD groups. We speculate that additional inputs from the cerebellum could overcome impaired information processing by favoring internal movement timing (Drucker et al., 2019). The efficacy of an online compensatory role of the cerebellum (Hanakawa et al., 1999; Drucker et al., 2019) is suggested in our study by the relatively preserved SCoM temporal movement sequencing (Avanzino et al., 2016), which could have also prevented the appearance of any gait freezing episode during our acquisitions. Relative timing of segmental movements was also described as unaltered in patients with PD by Rosin and colleagues (1997), further suggesting a compensatory rather than detrimental role of the cerebellum in parkinsonian patients with FOG and balance disturbances (Richard et al., 2017; Drucker et al., 2019; Isaias et al., 2020). Of relevance, the high variability in the SCoM movement onsets might have prevented us to detect differences across groups. Further studies with larger cohorts might further explore this aspect, to definitively rule out the presence of PD-related alterations in the movement sequencing.

We envisioned a significant impact of postural abnormalities on GI in PD, but our results did not support this hypothesis. Interestingly, our findings confirmed instead previous physiological studies reporting no correlation between APAs execution at GI and the natural inclination of the trunk (Leteneur et al., 2013) or of a forward leaning up to 30% of the maximum voluntary lean (Fawver et al., 2018).

Our study suffers from some limitations. First, although we reduced as much as possible the influence of known confounders (i.e., initial feet position and posture, anthropometric parameters, and cues), we cannot fully exclude a residual influence of parkinsonian symptoms, such as bradykinesia and rigidity, on the task performance (Delafontaine et al., 2017). However, in our previous work (Palmisano et al., 2020a), we showed that levodopa intake, by improving bradykinesia and rigidity, increases the length and speed of the first step at GI, but it does not affect the AP shift during UNL. We can therefore hypothesize that the alterations in AP displacement during UNL in the PDF group are not related to akinetic-rigid symptoms, but to impairment of the motor program itself. Future studies are needed to better clarify this aspect. Second, the limited sample size and a very stringent statistics may have limited the detection of differences between groups (e.g., SCoM onset times). Third, the lack of a brain imaging evaluation in this study prevents any firm conclusions of our pathophysiological interpretation of the kinematic and dynamic findings, but they match well with the brain metabolic activity changes (Tard et al., 2015, Lhomond et al., 2019) and network derangements (Lipski et al., 2017; Arnulfo et al., 2018; Georgiades et al., 2019; Pozzi et al., 2019) described during actual gait and gait freezing episodes in parkinsonian patients.

In conclusion, our data demonstrate substantial impairment of feedforward motor programming mechanisms at GI in parkinsonian patients. The deterioration of the UNL and stepping in PDF patients would suggest in these patients an additional impaired integration of postural and locomotor programs subserving gait initiation and modulation, which might be partly compensated by cerebellar mechanisms triggering time-locked models of body movement. Postural alterations seem to play a minor role on GI impairment in patients with PD. Last but not least, our results suggest the clinical utility of recording the CoP displacement during GI, and particularly its AP shift during the UNL, to possibly identify patients at risk of FOG and to monitor the efficacy of therapeutic strategies. Future longitudinal studies may support this assumption.

## 5 Contribution to the field

People with Parkinson’s disease frequently experience gait freezing, a sudden episodic inability to produce effective stepping despite the intention to walk. This dramatic phenomenon frequently occurs at gait initiation and leads to frequent falls, loss of independence and poor quality of life. The limited knowledge of the pathophysiological mechanism underlying freezing of gait still prevents an adequate and effective treatment. Poor agreement in literature may rely in particular on the multiple and poorly standardized methods adopted to study this motor task.

In this paper, we propose a robust methodological approach that may help minimize the influence of confounding factors on biomechanical outcome measurements of gait initiation in parkinsonian patients.

Most of relevance, patients with a positive history of gait freezing showed a marked deterioration of anticipatory postural adjustments at gait initiation and alterations of the unloading phase. These findings suggest poor ability in programming the motor task and integrating somatosensory information at gait initiation. As the relative timing of segmental movement was preserved, we can further hypothesize some compensatory cerebellar role for the execution of this motor task. These results are of relevance in defining grounds for pathophysiological hypotheses on the origin of this elusive motor symptom and may help patient monitoring and the development of new physiotherapeutic approaches specific for gait freezing.

## Supporting information

STOBE checklist

## Data Availability

All data produced in the present study are available upon reasonable request to the authors

## 7 Author Contributions

Chiara Palmisano: Conceptualization, Methodology, Software, Formal analysis, Investigation, Data curation, Writing – original draft. Laura Beccaria: Formal analysis, Investigation, Writing – original draft. Stefan Haufe: Formal analysis, Writing – review & editing. Jens Volkmann: Writing – review & editing. Gianni Pezzoli: Writing – review & editing, Funding acquisition. Ioannis U. Isaias: Conceptualization, Methodology, Formal analysis, Investigation, Resources, Writing – review & editing, Supervision, Project administration, Funding acquisition.

## 9 Acknowledgments

We would like to thank all patients and caregivers for their participation. Our special thanks go to Prof. Monica Norcini for study management and administrative support. The draft manuscript was edited for English language by Deborah Nock (Medical WriteAway, Norwich, UK).

